# Identifying the modification factors of exercise therapy on biomechanical load in patients with knee osteoarthritis: A systematic review and meta-analysis

**DOI:** 10.1101/2022.05.14.22275072

**Authors:** Moeka Yokoyama, Hirotaka Iijima, Keisuke Kubota, Naohiko Kanemura

**Affiliations:** Sportology Center, Graduate School of Medicine, Juntendo University, Tokyo, Japan; Graduate Course of Health and Social Services, Graduate School of Saitama Prefectural University, Saitama, Japan; Graduate School of Medicine / Institute for Advanced Research, Nagoya University Nagoya, Japan; Research & Development Center, Saitama Prefectural University, Saitama, Japan; Department of Physical therapy, Health and Social Services, Saitama Prefectural University, Saitama, Japan

**Keywords:** systematic review, meta-analysis, knee osteoarthritis, exercise therapy, biomechanical loads

## Abstract

**Objective:** As the progression of knee osteoarthritis (OA) is associated with large biomechanical loads, an optimally designed intervention is needed to prevent disease progression and symptoms. We aimed to investigate the effect of exercise therapy—the gold standard intervention—on biomechanical loads in patients with knee OA and identify its modification factors.

**Design:** Systematic review and meta-analysis

**Data sources:** PubMed, PEDro, and CINAHL; from study inception to May 2021

**Eligibility criteria:** Studies evaluating the first peak knee adduction moment (KAM), peak knee flexion moment (KFM), maximal knee joint compression force (KCF), or co-contraction during walking before and after exercise therapy in patients with knee OA

**Risk of Bias:** PEDro scale and NIH scale.

**Results:** Among 11 RCTs and nine non-RCTs, 1100 patients with knee OA were included. First peak KAM (SMD 0.11; 95% CI: -0.03–0.24), peak KFM (SMD 0.13; 95% CI: -0.03–0.29), and maximal KCF (SMD 0.09; 95% CI -0.05–0.22) tended to increase. An increased first peak KAM was significantly associated with a larger improvement in knee muscle strength and WOMAC pain. The quality of evidence regarding the biomechanical loads was low-to-moderate according to the GRADE approach.

**Conclusions:** Exercise therapy tends to increase biomechanical loads. The improvement in pain and knee muscle strength may mediate the increase in first peak KAM, suggesting difficulty in balancing symptom relief and biomechanical load reduction. Therefore, exercise therapy may satisfy both aspects simultaneously when combined with biomechanical interventions, such as a valgus knee brace or insoles.

**Funding:** Grant-in-Aid for JSPS Research Fellows, 19J23020.

**Registration:** PROSPERO (CRD42021230966)

## 1. INTRODUCTION

Rehabilitation of patients with knee osteoarthritis (OA) is primarily conducted to reduce pain and improve activities of daily living; in particular, exercise therapy is strongly recommended as a means to achieve the abovementioned goals, owing to its efficacy and safety^1^. Conversely, considering that OA severity is associated with an increase in pain and dysfunction^2,3^, it is important to deal with both the factors of knee OA progression and current symptoms to prevent exacerbation in the future. Increased biomechanical loads—such as the knee adduction moment (KAM) and knee flexion moment (KFM)—are generally considered risk factors of progression based on the results of several longitudinal studies^4–6^. Therefore, attention has been paid to whether exercise therapy can also reduce biomechanical load in patients with knee OA and slow disease progression.

The effect of exercise therapy on biomechanical loads currently remains unclear; some studies have reported a significant reduction in biomechanical loading^7^ while others have reported no significant effect^8,9^ or increment^10^. Additionally, although two previous systematic reviews investigated the effect of exercise therapy on peak KAM, only a limited number of studies were included, as the purpose was to clarify the relationship between altered peak KAM, pain, and walking speed after exercise therapy^8,9^. Therefore, comprehensive knowledge is required regarding the effect of exercise therapy on biomechanical loads, which may provide scope for exercise therapy.

When designing exercise therapies, it is also important to consider modification factors (i.e., moderator and mediator variables) that may influence the effect of intervention on biomechanical loads in patients with knee OA. Baseline characteristics that interact with the intervention to influence outcomes are defined as moderators, which suggest to clinicians appropriate populations that might be most responsive to the intervention^11^. Mediators are factors that play an intermediary role in the link between treatment and outcome^12^ and help us to identify possible mechanisms through which a treatment might achieve its effects^11^. For example, a previous systematic review identified moderators of exercise therapy that had different effects on improving pain in patients with knee OA, depending on the degree of joint deformity^13^. If the differences in biomechanical load after exercise therapy are also based on differences regarding the change in physical characteristics (due to the intervention or characteristics in patients with knee OA) from the baseline, it may be possible by synthesizing previous research to address potential moderators and/or mediators for future study, or make decisions regarding interventions. Therefore, we conducted this systematic review and meta-analysis to clarify the effect of exercise therapy on knee joint biomechanical load in patients with knee OA and identify factors that influence the biomechanical effect of exercise therapy. This knowledge will serve as a framework for future studies designed to optimize exercise therapy to prevent the progression of knee OA.

## 2. METHODS

This review followed the recommendations and guidelines of the Preferred Reporting Items for Systematic Reviews and Meta-analysis^14^ and the implementing PRISMA in Exercise, Rehabilitation, Sport medicine, and SporTs science guidance^15^ (Supplementary Checklist). The protocol was registered in PROSPERO (CRD42021230966; https://www.crd.york.ac.uk/prospero/display_record.php?RecordID=230966).

### 2.1 Eligibility criteria

In the present systematic review, biomechanical loads—including peak KAM, peak KFM, knee joint compression force (KCF), and co-contraction around the knee joint during walking—were considered primary outcomes, as these variables were reported to indicate knee OA progression^4,6,16,17^. We included studies that compared the primary outcomes at both pre- and postexercise therapy in patients diagnosed with tibiofemoral knee OA (unilateral and/or bilateral). All studies were published in peer-reviewed journals and written in English.

### 2.2 Literature search

Three electronic journal databases were searched from study inception to May 2021: PubMed, the Physiotherapy Evidence Database (PEDro), and the Cochrane Central Register of Controlled Trials. The specific search strategies are shown in the Supplementary Method. Google Scholar functioned as a complementary search engine, and a manual search of the reference lists of previous systematic reviews^8,9^ was also performed. After excluding duplicate studies, two reviewers (MY and HI) independently evaluated all titles and abstracts to determine whether they would be eligible for subsequent full-text review. The same reviewers independently reviewed the full-text articles for eligibility, and disagreements were resolved by discussion to reach a consensus.

### 2.3 Data extraction

A single reviewer (MY) collected the data, including the names of the authors, year of publication, patient population, knee OA severity, primary outcomes, potential moderator and mediator candidates, and funding sources; the same reviewer confirmed the collected data twice. One of the primary outcomes—the KAM—is known to have a bimodal waveform during the stance phase. Generally, the overall peak occurs at the first peak KAM; however, there is a possibility that the obtained peak values occur in different phases before and after intervention. To avoid this possibility, we contacted to the authors of the study to obtain the mean and standard deviation (SD) for the first peak KAM; if it could not be obtained, we used the peak KAM under the assumption that the peak KAM has a similar trend to the first peak KAM after exercise therapy. Potential moderator candidates included OA severity and joint deformity at the baseline. The potential mediator candidates included the standardized mean differences (SMDs) of body properties such as muscle strength and joint range of motion, as studies have hypothesized that biomechanical load reduction can be achieved through muscle strengthening^18,19^. Additionally, Harato et al. reported that in healthy adults, limiting knee extension by wearing orthoses increased peak KFM^20^. Although the SMDs of walking speed and pain were not pre-incorporated as potential mediators in our systematic review protocol in PROSPERO, we included these variables in our study as they may increase the amplitude of the peak KAM^10,21^. Unless otherwise indicated, the data of limbs with the most severe OA were used; if the mean and SD could not be extracted from the paper, we contacted the author of the study; in cases where no response was received from the authors or where they could not be contacted, we tried to obtain the data by extraction from figures using WebPlotDegitizer (https://automeris.io/WebPlotDigitizer/) software, converting the 95% confidence interval (CI) into the SD value using RevMan calculator, or converting the baseline and changed values after exercise therapy into mean and SD values using RevMan calculator. We also made contact with the authors to obtain data regarding the potential moderator and mediator candidates when they did not list them in their paper.

### 2.4 Assessing the risk of bias of the included studies and level of evidence of the outcomes

The risk of bias was independently assessed by two reviewers (MY and HI), using the PEDro scale^22^ for RCTs, and the NIH quality assessment tool for before-after (Pre-Post) studies with no control group scale^23^ for non-RCTs. The PEDro scale, which has 11 questions, was assessed as high quality when the score was more than six points (eight points are optimal for an exercise therapy study) ^24^. NIH scores—which include multiple questions assessing the potential risk for selection bias, information bias, measurement bias, and confounding—were classified as “good,” “fair,” or “poor” quality. Where there was disagreement between the reviewers, a consensus was reached through discussion.

### 2.5 Data analysis

Regardless of the intervention period or combination of other interventions, we conducted a meta-analysis of the primary outcomes. The pooled estimate and 95% CIs for SMDs were calculated using Review Manager version 5.4 (Cochrane Collaboration, London, United Kingdom). We used SMDs of the first peak KAM, peak KFM, and maximal KCF, because several studies did not report the weight or height of patients with knee OA, and the units could not be standardized^7,10,25–30^. Regarding maximal KCF, Messier et al.^31^ combined exercise therapy with dietary intervention; in our study, we divided the mean and SD values by the average patient weight before and after intervention to minimize the effect of combined diet therapy.

Meta-analysis was performed using a random-effects model^32^. To clarify the effect of exercise therapy on biomechanical loads in patients with knee OA, we analyzed within-group differences in biomechanical loads. The size of the SMD was interpreted using Cohen’s d (<0.5: small effect size; 0.5–0.8: moderate effect size; ≥0.8: large effect size)^33^. There are several methods to synthesize mediation effects in a systematic review, which is use of mediation effects reported in each study, or conduct a meta-mediation analysis in studies that included non-exercise and exercise groups. However, no studies have reported the mediation effects of exercise therapy on biomechanical load, and only a few studies allocated both non-exercise and exercise groups^25,27,28,34,35^. Therefore, we conducted regression analyses for the SMD size of the biomechanical loads in each study using the potential moderator candidate data at the baseline and the SMD size of each mediator as explanatory variables. If the number of corresponding data points was less than three, no regression analysis was performed. Statistical analyses were conducted using MATLAB 2020b (MathWorks Inc., Natick, MA, USA).

### 2.6 Quality assessment of the body of evidence: GRADE approach

The quality of evidence within each study was assessed using the Grading of Recommendations Assessment, Development, and Evaluation (GRADE) approach^36^. The quality of the body of evidence for each primary outcome measure was downgraded by criteria based on five domains (risk of bias, inconsistency, indirectness, imprecision, and publication bias) ^36^. The interstudy heterogeneity of the synthesized data was assessed using *I*^2^, which can be used even when the number of included studies is small^37^. A funnel plot was generated to assess publication bias, and Egger’s test^38^ for funnel plot asymmetry was performed in cases where the number of studies in each outcome measure was >10; if the number was ≤10, we assessed funnel plot asymmetry visually.

## 3. RESULTS

### 3.1 Study selection and characteristics of the included studies

Figure 1 shows a flowchart of the study selection process. Among a total of 776 articles, 62 were selected for full-text review, among which 41 were excluded as shown in Supplementary Table 1. Two studies by Foroughi et al. (2011a^39^ and 2011b^40^) met the inclusion criteria; however, they seemed to have used some of the same subjects; therefore, one study (Foroughi et al., 2011b) was excluded to avoid the duplication of patients.

**Figure 1.**
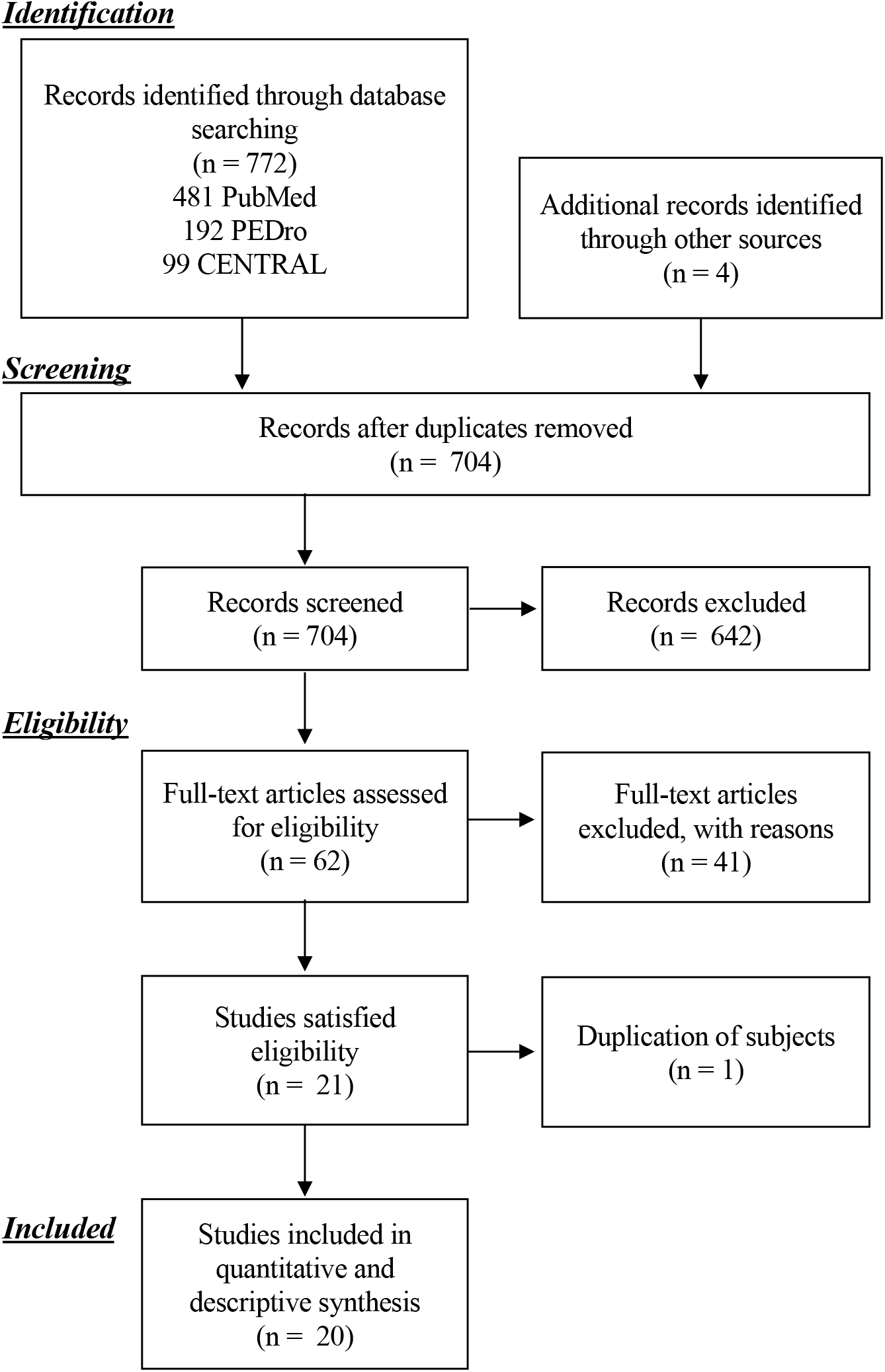
PRISMA flowchart.

In total, 1100 patients (average age: 63.8 years; Supplementary Table 2) from 11 RCTs^10,25,27–29,31,34,35,39,41,42^ and nine non-RCTs^7,26,30,43–48^ were included in the meta-analysis. Seven studies reported knee OA severity using the Kellgren and Lawrence grade^49^ or Outerbridge classification^50^. Supplementary Table 3 summarizes the intervention protocols used in the included studies. The median duration of the interventions was 12 weeks (range, 4 weeks to 18 months). The types of interventions included muscle strengthening, circuit training, neuromuscular training, proprioceptive training, stretching, massage, and balance training.

### 3.2 Effect of exercise therapy on the biomechanical loads

The first peak KAM (15 studies; n=449), peak KFM (eight studies; n=319), and maximal KCF (three studies; n=570) were synthesized by meta-analysis, which indicated no statistically significant difference in the effects of exercise therapy on first peak KAM (0.11; 95% CI: -0.03–0.24; Figure 2A), peak KFM (0.13; 95% CI: -0.03– 0.29; Figure 2B), or maximal KCF (0.09; 95% CI: -0.05–0.22; Figure 2C) during walking; instead, these outcome variables tended to increase postintervention. Due to the limited number of included studies, co-contraction around the knee joint was not included in the meta-analysis. Al-Khlaifat et al. reported a decrease in co-contraction between the vastus lateralis and biceps femoris during walking at the early and mid-stance phases following six weeks of exercise therapy, even if peak KAM did not significantly change during the same phases^43^.

**Figure 2.**
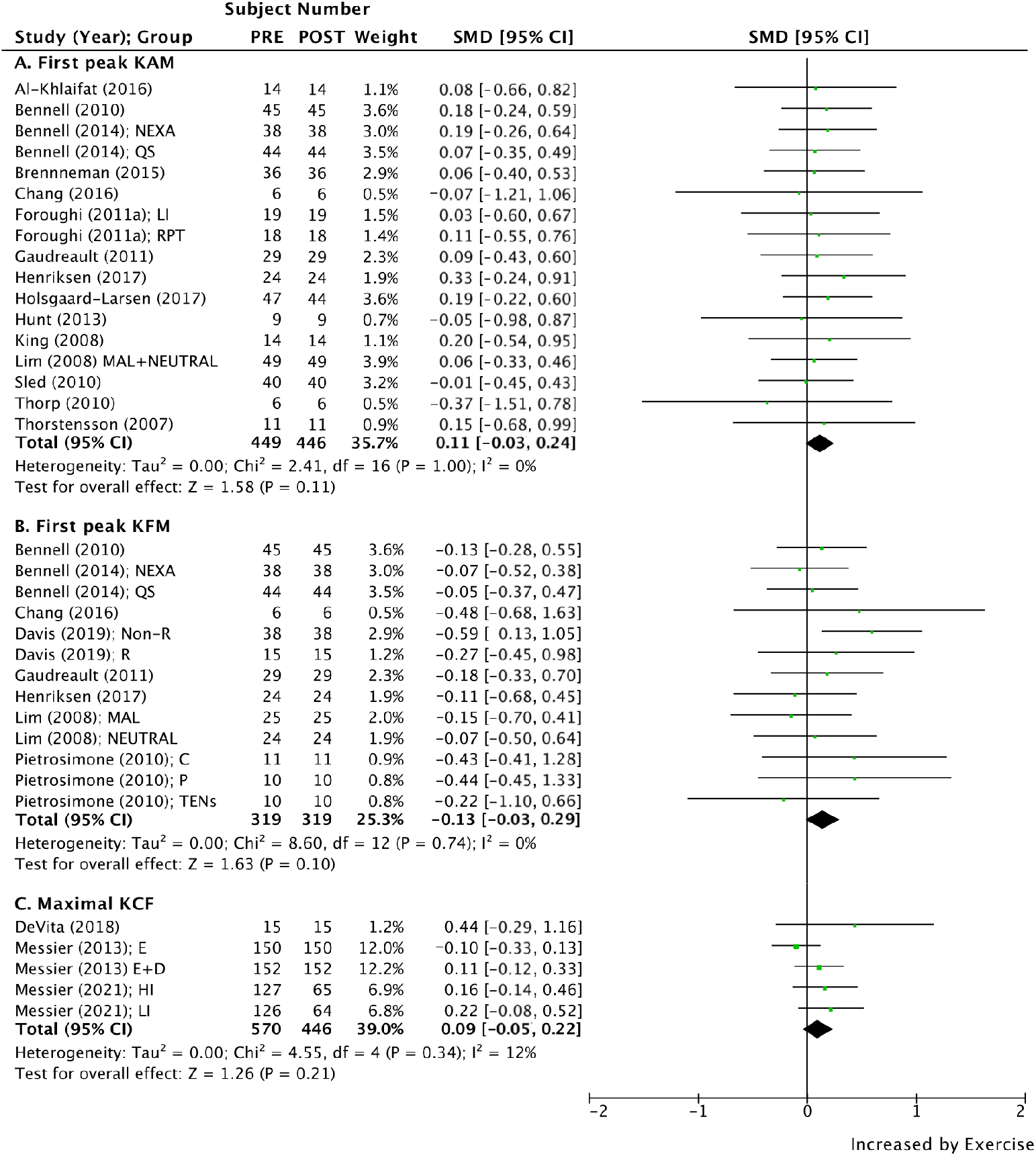
Standardized mean difference (SMD) and 95% confidence interval (CI) for the effect of exercise interventions on knee biomechanical load. (A) First peak KAM, (B) peak KFM, and (C) maximal KCF. Black diamonds represent the pooled effect size. The solid vertical line at 0 represents no significant difference.

### 3.3 Potential moderators and mediators that influence the effect of exercise therapy on biomechanical loads

Data of the following potential moderator and mediator candidates were obtained (Figure 3): OA severity, femoral-tibia angle (FTA), hip-knee-ankle angle (HKA) (Supplementary Table 1), SMDs of muscle strength (Supplementary Figures 1–2), pain (Supplementary Figure 3A), and walking speed (Supplementary Figure 3B). Additionally, the authors of several studies kindly provided unpublished data on biomechanical loads and potential moderator and mediator candidates (see details in Supplementary Table 4); however, because no study reported the range of motion or extensibility of the muscle of interest (some studies included a muscle stretch session in their intervention protocol), these variables were not assessed in the present study.

**Figure 3.**
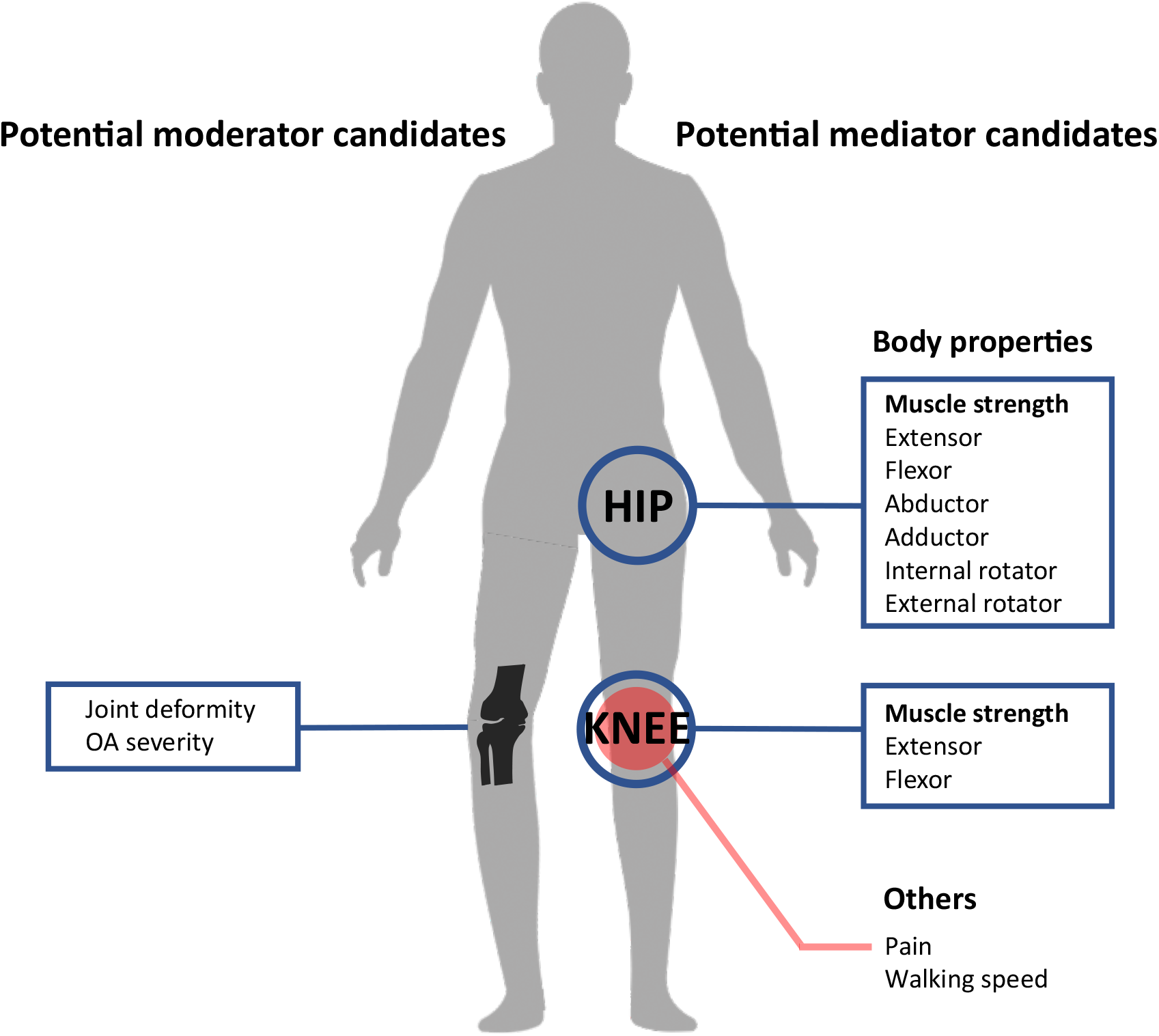
Potential moderator and mediator candidates.

Figure 4 and Supplementary Figures 4 and 5 show the relationship between the SMDs of the biomechanical loads and the potential moderator candidates or those of mediator candidates after exercise therapy. We also confirmed a significant relationship between an increase in peak KAM and improvement in knee extensor muscle strength (β coefficient: 0.35, p=0.010; Figure 4A), knee flexor muscle strength (β coefficient: 0.57, p=0.035; Figure 4B), and Western Ontario and McMaster Universities Osteoarthritis Index (WOMAC) pain (β coefficient: -0.26, p=0.041; Figure 4C). We also found that an increase in the first peak KAM was associated with a slower walking speed (β coefficient: -0.42, p=0.020; Supplementary Figure 5A); however, no relationship was found between peak KFM and maximal KCF force with potential moderator (Supplementary Figure 4B) or mediator (Supplementary Figure 5 B, C) candidates.

**Figure 4.**
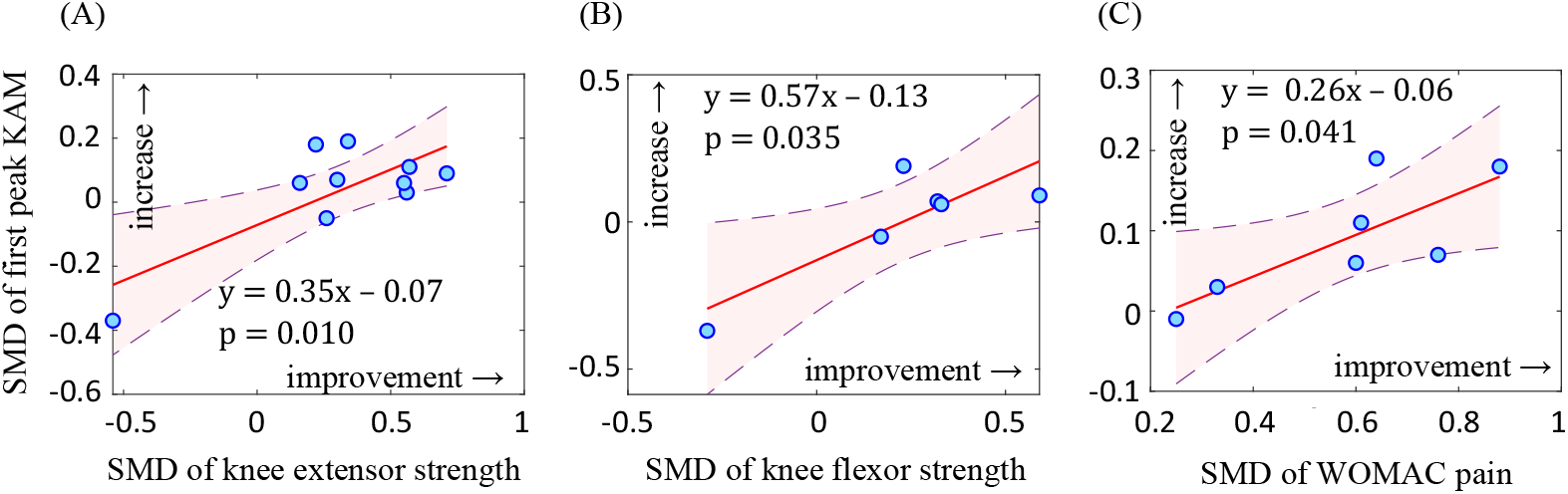
Factors that increased the first peak KAM. The red solid line represents the regression line, and the purple dashed line indicates the 95% confidence interval of the regression line.

### 3.4 Risk of bias

The risk of bias for each study is shown in Supplementary Tables 5–6. Ten of 11 RCTs demonstrated high quality data (average: 6.8 points) when using the PEDro scale; two non-RCTs demonstrated good quality, and seven non-RCTs demonstrated fair quality (average: 6.1 points) data. The funnel plots used to assess publication bias are shown in Supplementary Figure 6; no publication bias was observed for the first peak KAM or peak KFM in Egger’s test, or for maximal KCF in visual assessment.

The results of the GRADE approach are shown in Table 1. The first peak KAM and peak KFM started with a low-quality rating, as 53% and 37% of the studies included in each outcome were non-RCTs, respectively. The maximal KCF started with a high-quality rating, as all included studies in the present systematic review were RCTs. However, the evidence of these outcomes were downgraded due to imprecision; thus, the first peak KAM and peak KFM were rated as low-quality, and the maximal KCF quality rating was moderate.

**Table 1.**
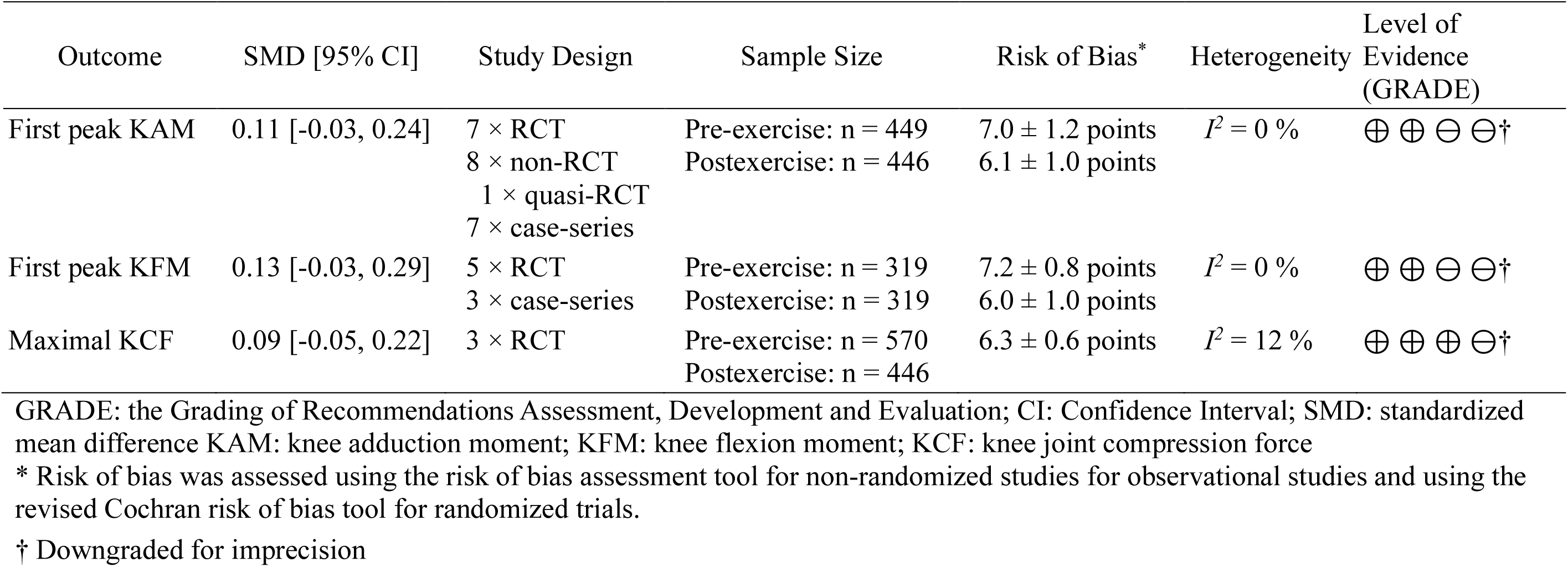
Summary of the body evidence according to the GRADE approach

## 4. DISCUSSION

Previous studies prescribed exercise therapy to patients with knee OA under the assumption that biomechanical loads would be reduced through muscle strengthening. However, the effect of exercise therapy on the biomechanical load, and modification factors that could explain the variation in the biomechanical load results, remain unclear. We performed a meta-analysis of 11 RCTs and nine non-RCTs that investigated the biomechanical effects of exercise therapy among 1100 patients with knee OA. Contrary to the biomechanical load reduction theory^18,19^, our study revealed that chronic exercise tended to increase the first peak KAM, peak KFM, and maximal KCF when compared with the baseline values. Exercise therapy improved periarticular knee muscle strength and pain, which are potential factors mediating the increased first peak KAM. This suggests that managing improvements in pain or physical function and biomechanical load reduction—especially the first peak KAM—is difficult when using current exercise therapy, possibly due to the simultaneous improvement in knee pain and muscle strength.

### 4.1 Potential modification factors for the effect of exercise therapy on first peak KAM

Body properties—particularly muscle strength—have frequently been targeted for exercise therapy. Most of the studies included in the present systematic review targeted strengthening of the hip abductor and knee extensor muscles for the following reasons: hip abductor muscle weakness is considered to increase KAM by keeping the center of mass (COM) away from the knee joint by enhancing pelvic contralateral tilt,^19^ while knee extensors are thought to decrease KAM by acting as knee stabilizers in the frontal plane^18^. Contrasting with the proposed mechanism, our results revealed an increase in first peak KAM as knee extensor muscle strength increased, despite the reports of several studies that combined the strengthening of hip abductor muscles^7,25,26,28,30,35,39,41,43,48^. For example, Bennell et al. performed exercise therapy for 12 weeks, targeting hip abductors and knee extensors; they demonstrated increased pelvic contralateral tilt and decreased trunk ipsilateral lean^25^. As this altered motion after exercise therapy followed the increased KAM mechanism that keeps COM away from the knee joint center, it is possible that these characteristic movement patterns in patients with knee OA were emphasized through increased muscle strength. Interestingly, the first peak KAM did not increase with walking speed after exercise therapy; conversely, a previous systematic review suggested that walking speed affected the amplitude of joint moment in patients with knee OA, as well as in different populations^51^. Considering results from our regression analysis and a previous systematic review that suggested resistance training would enhance walking speed, but not KAM, in patients with knee OA^9^, it may be suggested that patients with knee OA demonstrate an increase in the first peak KAM through different pathways than the increase in the first peak KAM owing to an increase in walking speed. Other potential mediators, such as pain improvement, also increased the first peak KAM. This effect has been demonstrated after pharmacotherapy, including intra-articular injection of hyaluronic acid^52^ and pain relief medication for patients with knee OA^53^. Additionally, a relationship between pain and peak KAM has also been demonstrated in healthy young adults. Henriksen et al. reported that intra-articular injection of hypertonic saline decreased peak KAM and increased pain^54^. Pain improvement may therefore modify the effect of intervention on the first peak KAM; however, it cannot be denied that the increase in biomechanical load in patients with knee OA was caused by muscle strengthening following pain relief, as the first peak KAM did not significantly change after intra-articular injection of lidocaine, a local anesthetic^55^. To summarize these results, it should be noted that pain relief and the reduction of biomechanical loads were not simultaneously satisfied using exercise therapy or pharmacotherapy alone. This systematic review did not identify any factors associated with peak KFM or maximal KCF after exercise therapy in patients with knee OA, suggesting that other moderator or mediator candidates that were not selected for this study may exist. For example, Harato et al. reported knee extension limitations in healthy adults when wearing orthoses, which increased the peak KFM^20^. Campbell et al. also reported that flexion contracture at the knee joint was associated with disease progression and clinical outcomes;^56^ therefore, mobility at the knee joint may be a potential moderator or mediator candidate for future studies.

### 4.2 Suggestion for future research and intervention

Muscle strengthening is a common and effective intervention to improve pain and physical function in daily life among patients with knee OA^57^. It is strongly recommended due to its low risk and high cost-effectiveness^1^; however, we revealed a potential disadvantage of exercise therapy, which slightly affects the increment in biomechanical loads. As we were not able to reveal effective targets to reduce biomechanical loads in the present systematic review, exercise therapy needs to be considered in order to balance benefits and risks until effective targets are clarified. According to the results of our regression analysis, this systematic review revealed that the increase in peak KAM depends on the amount of improvement in the knee extensor or flexor muscles. However, an easy interpretation that makes adjustment for muscle strengthening—especially of the periarticular knee muscle—should be avoided. Hall et al. confirmed that every 1-unit [Nm/kg] increase in knee extensor strength was associated with a physical function improvement of 17 WOMAC units^58^. This could therefore undermine the benefits of muscle strengthening on physical function. Based on the results of our study, we proposed another option that combines exercise therapy with other therapies that have effects on biomechanical load reduction. Previous systematic reviews demonstrated a significant reduction in the first peak KAM of both valgus knee brace (effect size: 0.61; 95% CI: 0.39–0.83)^59^ and insole (effect size: 0.23; 95% CI: 0.07–0.40) interventions^60^. To the best of our knowledge, no studies have investigated the effect of exercise combined with these interventions on biomechanical loads; however, it may counteract the increase in peak KAM after exercise therapy. Therefore, in the future, we need to identify a combination of interventions that maximizes the effect on both symptoms and biomechanical loads.

### 4.3 Limitations

One of the methodological limitations of the present systematic review is that it was not the most appropriate way to investigate mediators. We conducted regression analyses as an alternative means of mediator analysis, as no study reported the mediation effect of exercise therapy on biomechanical load; only a few studies adopted an RCT design that allocated both a non-exercise and exercise group. The lack of RCTs that allocated both non-exercise and exercise groups also downgraded the quality of evidence regarding primary outcomes in the systematic review. Therefore, further RCTs are needed to provide robust evidence on the effect of exercise therapy on biomechanical load, and to confirm the relationship between suggested mediators and biomechanical load.

## 5. CONCLUSION

To summarize the results of the present systematic review, exercise therapy in patients with knee OA tended to favor increased biomechanical loads, including first peak KAM, peak KFM, and maximal KCF. Based on the regression results, this tendency seems to be a result of relieving knee joint pain and improving muscle strength in knee extensors and flexors, especially regarding the peak KAM. Since we could not identify an effective intervention target to reduce biomechanical loads, further exercise therapy studies with other intervention perspectives are required. Our results also suggest the necessity of future studies to identify the proper intensity of muscle strengthening for the knee muscle, and the combination of interventions maximizing the effect on both symptoms and biomechanical load.

## Data Availability

All data produced in the present work are contained in the manuscript

## Acknowledgement

This research was supported by a Grant-in-Aid for JSPS Research Fellows (grant number: 19J23020).

## Contributions

All authors have read and approved the submitted manuscript.

Study design: Moeka Yokoyama, Hirotaka Iijima, Naohiko Kanemura

Article collection: Moeka Yokoyama, Hirotaka Iijima, Keisuke Kubota

Main reviewer: Moeka Yokoyama, Hirotaka Iijima

Meta-analysis: Moeka Yokoyama

Manuscript composition: Moeka Yokoyama, Hirotaka Iijima, Keisuke Kubota, Naohiko Kanemura

