## Supplementary Materials for "Identifying the modification factors of exercise therapy on biomechanical load in patients with knee osteoarthritis: A systematic review and meta-analysis"

**Table of contents**

**1. Supplemental Method** – Search strategies (Page S2)

**2. Supplemental Table**

**Supplemental Table 1.** Reason for exclusion (Page S3)

**Supplemental Table 2.** Characteristics of study (Page S4-S8)

**Supplemental Table 3.** Summary of the intervention protocols (Page S9-S12)

**Supplemental Table 4.** List of variables of biomechanical loads and potential moderator and mediator candidates (Page S13)

**Supplemental Table 5.** Risk of bias in randomized controlled trials (Page S14)

**Supplemental Table 6.** Risk of bias in non-randomized controlled trials (Page S15)

**3. Supplemental Figure**

**Supplemental Figure 1.** SMD and 95% CI for muscle strength of knee extensor and knee flexor in each group (Page S16)

**Supplemental Figure 2.** SMD and 95% CI for muscle strength of hip extensor , hip flexor , hip abductor , hip adductor , hip external rotator, and hip internal rotator in each group (Page S17)

**Supplemental Figure 3.** SMD and 95% CI for WOMAC pain (A) and walking speed (B) in each group (Page S18)

**Supplemental Figure 4.** Relationship between SMD for first peak KAM or first peak KFM and potential moderator candidate (Page S19)

**Supplemental Figure 5.** Relationship between SMD for first peak KAM or first peak KFM, or maximal KCF and potential mediator candidates (Page S20)

**Supplemental Figure 6.** Funnel plot representing publication bias for the effect of exercise intervention on first peak KAM, peak KFM, and maximal KCF (Page S21)

**4. References**

**Supplementary Method.** Search strategy

PubMed (within all fields)

#1 “knee joint” [MeSH] OR knee

#2 “Osteoarthritis” [MeSH]

#3 “Walking” [MeSH]

#4 “biomechanical phenomena” [MeSH] OR “biomechanics” OR “moment” OR “compression”

#5 “electromyography” OR “EMG”

#6 “Physical Therapy Modalities” [Mesh] OR “Exercise” [Mesh]

#7 #1 AND #2

#8 #4 OR #5

#9 #7 AND #3 AND # 8 AND #6

PEDro

#1 knee, osteoarthritis, gait [within following fields: Therapy: strength training; Method: clinical trial]

#2 knee, osteoarthritis, gait [within following fields: Therapy: stretching, mobilisation, manipulation, massage; Method: clinical trial]

#3 knee, osteoarthritis, walking [within following fields: Therapy: strength training; Method: clinical trial]

#4 knee, osteoarthritis, walking [within following fields: Therapy: stretching, mobilisation, manipulation, massage; Method: clinical trial]

CENTRAL (within trial)

#1 knee

#2 osteoarthritis

#3 “walking”

#4 “biomechanical phenomena” or “biomechanics” or “moment” or “compression”

#5 “electromyography” or “EMG”

#6 “physical therapy modalities” or “exercise”

#7 #1 and #2

#8 #4 or #5

#9 #7 and #3 and # 8 and #6

**Supplementary Table 1.** Reason for exclusion

| 1: No exercise intervention  2: No results for biomechanical loads (i.e. peak KAM, peak KFM, and maximal knee joint compression force at pre-/post-intervention  3: Patients either did not have knee OA or had posttraumatic knee OA | | | | | | | |
| --- | --- | --- | --- | --- | --- | --- | --- |
| **Author (year)** | **１** | **2** | **3** | **Author (year)** | **1** | **2** | **3** |
| Alkhawajah et al. (2019)^1^ |  | **●** |  | Hall et al. (2018)^2^ |  | **●** |  |
| Altmis et al. (2018)^3^ |  | **●** |  | Hanada et al. (2018)^4^ |  | **●** |  |
| Aoki et al. (2009)^5^ |  | **●** |  | Huang et al. (2005)^6^ |  | **●** |  |
| Bechard et al. (2012)^7^ |  | **●** |  | Ikuta et al. (2020)^8^ |  | **●** |  |
| Bennell et al. (2007)^9^ | **●** |  |  | Inal et al. (2016) ^10^ |  | **●** |  |
| Bennell et al. (2015)^11^ |  | **●** |  | Jafarnezhadgero et al. (2020) ^12^ |  |  | **●** |
| Brenneman (2018) ^13^ |  | **●** |  | Jan et al. (2009)^14^ |  | **●** |  |
| Capin et al. (2018)^15^ |  |  | **●** | Kean et al. (2017)^16^ |  | **●** |  |
| Cheing et al. (2004)^17^ |  | **●** |  | Kobsar et al. (2015)^18^ |  | **●** |  |
| Clausen et al. (2014)^19^ | **●** |  |  | Kobsar et al. (2017)^20^ |  | **●** |  |
| de Rooij et al. (2017)^21^ |  | **●** |  | Liang et al. (2019)^22^ |  | **●** |  |
| Drexler et al. (2012)^23^ | **●** |  |  | Lim et al. (2009) ^24^ | **●** |  |  |
| Durmus et al. (2007)^25^ |  | **●** |  | McCarthy et al. (2004)^26^ |  | **●** |  |
| Ebnezar et al. (2012)^27^ |  | **●** |  | Messier et al. (1997)^28^ |  | **●** |  |
| Elbaz et al. (2014)^29^ | **●** |  |  | Messier et al. (2000)^30^ |  | **●** |  |
| Farrokhi et al. (2017)^31^ | **●** |  |  | Mihalko et al. (2019)^32^ |  | **●** |  |
| Fransen et al. (2001)^33^ |  | **●** |  | Olagbegi et al. (2016)^34^ |  | **●** |  |
| Fisher et al. (1997) ^35^ |  | **●** |  | Peungsuwan et al. (2014)^36^ |  | **●** |  |
| Haim et al. (2012)^37^ | **●** |  |  | Rashid et al. (2019)^38^ |  | **●** |  |
| Hall et al. (2015)^39^ |  |  | **●** | Wang et al. (2018)^40^ |  | **●** |  |
| Total:40 | | | | | **7** | **30** | **3** |

**Supplementary Table 2.** Characteristics of study

| Author (Year) | Study Design | Number of Patients (Allocated/Analyzed) | Patient Population | Funding Source |
| --- | --- | --- | --- | --- |
| Al-Khlaifat et al. (2016) ^41^ | Pre/posttest | 19/14 | (Analyzed Population)  Age: 61.79 ± 10.42 y; height: 1.62 ± 0.08 m; body mass: 81.07 ± 15.82 kg; BMI: 30.89 ± 5.05 kg/m^2^; female: 86%; OA grade:-; HKA: -° | The University of Jordan and the University of Salford (ISRCTN No. 61720526) |
| Bennell et al. (2010) ^42^ | RCT | 45/39 | (Allocated Population)  Age: 64.5 ± 9.1 y; height: - m; body mass: - kg; BMI: 27.5 ± 4.7 kg/m^2^; female: 51%; OA grade: KL II (n = 15), III (n = 15), Ⅳ(n = 15) ;HKA: -°  (Analyzed Population)^*^  OA grade: KL II (n = 14), III (n = 13), Ⅳ(n = 12); HKA: 178.1 ± 3.2° | The National Health and Medical Research Council (Project #454686) |
| Bennell et al. (2014) ^43^ | RCT | E1: 50/38  E2: 50/44 | E1: NEXA (Neuromuscular exercise group)  (Allocated Population)  Age: 62.7 ± 7.3 y; height: 1.68 ± 0.09 m; body mass: 83.8 ± 13.5 kg; BMI: 29.6 ± 3.9 kg/m^2^; female: 52%; OA grade: KL II (n = 9), III (n = 21), Ⅳ(n = 20); HKA: 177.3 ± 3.0°  (Analyzed Population)  OA grade: KL II (n = 6), III (n = 17), Ⅳ(n = 15); HKA: 177.1 ± 2.9°  E2:QS (Quadriceps strengthening group)  (Allocated Population)  Age: 62.2 ± 7.4 y; height: 1.66 ± 0.10 m; body mass: 81.6 ± 15.1 kg; BMI: 29.7 ± 4.3 kg/m^2^; female: 52%; OA grade: II (n = 13), III (n = 22), Ⅳ(n = 15); HKA: 176.4 ± 3.9°  (Analyzed Population)  OA grade: KL II (n = 11), III (n = 21), Ⅳ(n = 12); HKA: 176.2 ± 3.9° | The National Health and Medical Research Council (project grant 628644; Fellowship 1002190).  The Australian Research Council (Future Fellowship FT0991413) |
| Brenneman et al. (2015) ^44^ | Pre/posttest | 45/36 | (Analyzed population)  Age: 60.3 ± 6.5 y; height: 1.63 ± 0.06 m; body mass: 78.1 ± 14.8 kg; BMI: 29.5 ± 5.3 kg/m^2^; female: 100%; OA grade: -; HKA: -° | Labarge Optimal Aging Initiative-Opportunities Fund (MRM)  Natural Sciences and Engineering Research Council of Canada-Discovery (#353715; MRM)  Canadian Foundation of Innovation and Ontario Ministry of Research and Innovation-Leaders Opportunities Fund (#27501; MRM) |

RCT: randomized controlled trial; BMI: body mass index; OA: osteoarthritis; KL: Kellgren and Lawrence
(Continued)

| Author (Year) | Study Design | Number of Patients (Allocated/Analyzed) | Patient Population | Funding Source |
| --- | --- | --- | --- | --- |
| Chang et al. (2016)^45^ | Pre/posttest | -/6 | (Analyzed population)  Age: - y; height: - m; body mass: - kg; BMI: -kg/m^2^; female: 100%; OA grade: -; HKA: -° | Memorial Hospital for the financial support (Grant no. SKH-8302-101-NDR-05). |
| Davis et al. (2019)^46^ | Pre/posttest | Overall  53/53  E1: 38  E2: 15 | (Allocated and analyzed population)  E1: Non-Responder group  Age: 62.89 ± 7.53 y; height: - m; body mass: 83.06 ± 13.66 kg; BMI: 28.14 ± 4.26 kg/m^2^; female: 55%; OA grade: KL II (n = 12), III (n = 20), IV (n = 6) ; HKA: -°  (Allocated and analyzed population)  E2: Responder group  Age: 61.13 ± 5.84 y; height: - m; body mass: 91.97 ± 18.23 kg; BMI: 29.33 ± 3.82 kg/m^2^; female: 40%; OA grade: KL II (n = 5), III (n = 9), IV (n = 1) ; HKA: -° | The National Institute of Arthritis and Musculoskeletal and Skin Diseases of the National Institutes of Health (1R21AR067560-01) |
| DeVita et al. (2018)^47^ | Two center RCT | E1: 16/15 | (Analyzed population)  Age: 58.1 ± 6.5 y; height: 1.73 ± 0.07 m; body mass: 79.4 ± 14.8 kg; BMI: 26.4 ± 4.0 kg/m^2^; female: 67%; OA grade: KL Ⅰ (n=2), II (n = 4), III (n = 7), IV (n = 2) ; HKA: - ° | The Biomechanics Laboratory in the Department of Kinesiology at East Carolina University  The Danish Rheumatism Association and The Oak Foundation |
| Foroughi et al.(2011a) ^48^ | RCT | E1: 28/19  E2: 26/18 | (Allocated population)  E1: Sham (Low-intensity training group) Age: 64 ± 8 y; height: 1.6 ± 6 m; body mass: 86.5 ± 23 kg; BMI: 33.2 ± 8.1 kg/m^2^; female: 100%; OA grade: mild (n=12), moderate (n=3), severe (n=7), very severe (n=6) ; HKA: -°  (Allocated population)  E2: PRT (Progression resistance training group) Age: 64 ± 7 y; height: 1.6 ± 6 m; body mass:82.0 ± 14 kg; BMI: 31.9 ± 5.2 kg/m^2^; female: 100%; OA grade: mild (n=6), moderate (n=5), severe (n=5), very severe (n=10) ; HKA: -° | The University of Sydney R & D Grant (S4201 U3301) |

RCT: randomized controlled trial; BMI: body mass index; OA: osteoarthritis; KL: Kellgren and Lawrence

(Continued)

| Author (Year) | Study Design | Number of Patients (Allocated/Analyzed) | Patient Population | Funding Source |
| --- | --- | --- | --- | --- |
| Gaudreault et al. (2011) ^49^ | Pre/posttest | 29/29 | (Allocated and analyzed population)  Age: 63.3 ± 8.4 y; height: 1.6 ± 0.1 m; body mass: 80.5 ± 18.3 kg; BMI: 31 ± 5 kg/m^2^; female: 76%; OA grade: KL I (n = 10), II (n = 5), III (n = 5), IV (n = 9); HKA: -° | Canadian Institute for Health Research (CIHR) grant  The CIHR Clinical Initiative Strategy program - postdoctoral fellowship award |
| Henriksen et al. (2016)^50^ | RCT | 31/24 | (Analyzed population)  Age: 64.9 ± 9.1 y; height: 1.69 ± 0.08 m; body mass: 83.1 ± 14.0 kg; BMI: 29.1 ± 4.1 kg/m^2^; female: 92%; OA grade: KL II (n = 15), III (n = 5), IV (n = 4); FTA: 25.8 ± 7.6° | - |
| Holsgaard-Larsen et al. (2017)^51^ | RCT | 47/44 | (Allocated population)  Age: 57.9 ± 7.9 y; height: - m; body mass: 79.1 ± 12.4kg; BMI: 26.8 ± 3.3 kg/m^2^; female: 61.7%; OA grade: KL I (n = 26), II (n = 14), III (n = 7) ; HKA: -° | The Region of Southern Denmark PhD Fund  The Region of Southern Denmark Research Fund  The Danish Rheumatism Association  The Danish Rheumatism Association Ryholts grant  The University of Southern Denmark Scholarship  The Association of Danish Physiotherapists  Odense University Hospital free research funds  Family Hede Nielsens Fund  The Oak Foundation |
| Hunt et al. (2013) ^52^ | RCT | 9/9 | (Allocated and analyzed population)  Age:- y; height: - m; body mass: - kg; BMI: -kg/m^2^; female: 44%; OA grade: KL II (n = 7), III (n = 2); HKA: 177.5 ± 2.8 ° | The Canadian Arthritis Network |

*1: The author represented distribution of OA severity as percentage, however, in total, it was 96.1%

RCT: randomized controlled trial; BMI: body mass index; OA: osteoarthritis; KL: Kellgren and Lawrence

(Continued)

| Author (Year) | Study Design | Number of Patients (Allocated/Analyzed) | Patient Population | Funding Source |
| --- | --- | --- | --- | --- |
| King (2008) ^53^ | Pre/posttest | 14/14 | (Allocated and analyzed population)  Age: 48.4 ± 6.5 y; height: 1.77 ± 0.08 m; body mass: 92.4 ± 14.2 kg; BMI: 29.3 ± 3.3 kg/m^2^; female: 14%; OA grade: KL I (n = 2), II (n = 4), III (n = 7), IV (n = 1); Mechanical axis: -6.9 ± 3.4° | The Canada Research Chairs Program (TBB); the Canadian Institutes of Health Research; the Arthrex Inc |
| Lim et al. (2008) ^54^ | RCT | E1: 26/25  E2: 27/24 | (Allocated population)  E1: MAL (more malaligned group) Age: 67.2 ± 6.7 y; height: 1.67 ± 0.10 m; body mass: 78.7 ± 13.0 kg; BMI: 28.2 ± 3.7 kg/m^2^; female: 50%; OA grade: KL II (n = 4), III (n = 8), IV (n = 14); Varus malalignment: 6.7 ± 1.7°(Analyzed population)  Age: 67.1 ± 6.8 y; female: 48%; OA grade: KL II (n = 3), III (n = 8), IV (n = 14); HKA: 174.1 ± 1.8°  (Allocated population)  E2: NEUTRAL (more neutral group) Age: 64.1 ± 9.3 y; height: 1.65 ± 0.1 m; body mass: 79.2 ± 15.9 kg; BMI: 29.0 ± 5.2 kg/m^2^; female: 63%; OA grade: KL II (n = 12), III (n = 7), IV (n = 8); Varus malalignment: 1.2 ± 1.9°  (Analyzed population)  Age: 64.3 ± 9.3 y; female: 62.5%; OA grade: KL II (n = 11), III (n = 6), IV (n = 7); HKA: 180.4 ± 2.6° | United Pacific Industries through a grant from the Physiotherapy Research Foundation |
| Messier et al. (2013)^55^ | RCT | E1:150/134  E2:152/136 | (Allocated population)  E1: E (exercise) group  Age: 66 ± 6 y; height: 1.66 ± 0.09 m; body mass: 93 ± 15.2 kg; BMI: 33.5 ± 3.7 kg/m^2^; female: 72%; OA grade: KL 2.53 ± 0.56 (overall); HKA: -°  (Allocated population)  E2: E (exercise) + D (diet) group  Age: 65 ± 6 y; height: 1.66 ± 0.09 m; body mass: 93 ± 14.4 kg; BMI: 33.6 ± 3.7 kg/m^2^; female: 72%; OA grade: KL 2.59 ± 0.60 (overall); HKA: -° | The National Institute of Arthritis and Musculoskeletal and Skin Diseases (R01 AR052528-01)  The National Institute on Aging (P30 AG21332)  The National Center for Research Resources (M01-RR00211)  General Nutrition Centers Inc, USA |

RCT: randomized controlled trial; BMI: body mass index; OA: osteoarthritis; KL: Kellgren and Lawrence

(Continued)

| Author (Year) | Study Design | Number of Patients (Allocated/Analyzed) | Patient Population | Funding Source |
| --- | --- | --- | --- | --- |
| Messier et al. (2021)^56^ | RCT | E1:126/108  E2:127/109 | (Allocated population)  E1: LI (low intensity) group  Age: 64 ± 8 y; height: 1.69 ± 0.10 m; body mass: 89± 18 kg; BMI: 31 ± 6 kg/m^2^; female: 41%; OA grade: KL II (n = 64), III (n = 48), IV (n = 14); HKA: -°  (Allocated population)  E2: HI (high intensity) group  Age: 67 ± 9 y; height: 1.68 ± 0.11 m; body mass: 89 ± 19 kg; BMI: 31 ± 6 kg/m^2^; female: 41%; OA grade: KL II (n = 63), III (n = 50), IV (n = 14); HKA: -° | The National Institute of Arthritis and Musculoskeletal and Skin Diseases (1R01AR059105-01)  The National Institute on Aging (P30 AG21332) |
| Pietrosimone et al. (2010) | RCT | E1:12/10  E2:12/10  E3:12/11 | E1:TENS; Age: - y; height: 1.71 ± 0.09 m; body mass: -; BMI: 28.6 ± 4.8 kg/m^2^; female: 50%; OA grade:3 ± 0.9; HKA: -°  E2:Placebo; Age: - y; height: 1.72 ± 0.08 m; body mass: -; BMI: 29.5 ± 9.8 kg/m^2^; female: 67%; OA grade:3.1 ± 0.8 ;HKA: -°  E3:Control;Age: - y; height: 1.70 ± 0.11 m; body mass: -; BMI: 28.6 ± 5.6 kg/m^2^; female: 58%; OA grade:2.9 ± 1.2; HKA: -° | The American Physical Therapy Association, Orthopedic Section Grant, National Athletic Trainers’ Association Research and Education Foundation, and EMPI Inc. |
| Sled et al. (2010) ^57^ | Pretest-posttest, control group | 40/40 | (Allocated and analyzed population)  Age: 62.98 ± 9.73 y; height: 1.73 ± 0.11 m; body mass: 82.31 ± 20.00 kg; BMI: 27.38 ± 5.47 kg/m^2^; female: 57%; OA grade: KL 2.5 ± 0.91 (overall) ; HKA: -° | Bickell Foundation of Canada Medical Research grant |
| Thorp et al. (2010) ^58^ | Pretest-posttest | 6/6 | (Allocated and analyzed population)  Age: 59.7 ± 17.2 y; height: - m; body mass: - kg; BMI: 30 ± 4.1kg/m^2^; female: 83%; OA grade: -; HKA: -° | - |
| Thorstensson et al. (2007) ^59^ | Pretest-posttest, | 13/11 | (Analyzed population)  Age: 54.6 ± 5.4 y; height: - m; body mass: - kg; BMI: 25.5 ± 2.6 kg/m^2^; female: 45%; OA grade: KL 0 (n = 2), I (n = 4), II (n = 4), III (n = 1); HKA: 182.2 ± 4.6° | The Norrbacka-Eugenia Foundation; The Halland County Council; The Swedish Research Council; The Swedish Rheumatism Association in Stockholm and Gothenburg; The Thelma Zoega Foundation; The Swedish National Centre for Research in Sports; Medical Faculty at Lund University |

RCT: randomized controlled trial; BMI: body mass index; OA: osteoarthritis; KL: Kellgren and Lawrence

**Supplementary Table 3.** Summary of the intervention protocols

| **Author (Year)** | **Frequency and Duration** | **Intensity** | **Contents of therapeutic exercise** |
| --- | --- | --- | --- |
| Al-Khlaifat et al. (2016) ^41^ | Once/week for 6 weeks; supervised  Once/day for 6 weeks; home-based | Three sets of each exercise at 40-50% of 10RM  (daily adjusted) | Circuit training  1. bilateral, split, and unilateral squats, step-ups, side lowers (with dumb-bells) with five levels of difficulty, side-lying hip abduction, clam, bridging, and knee extension exercises (with weight or TheraBand™).  2. cycling on a stationary bike  10-15 min exercise with weights and TheraBand™ |
| Bennell et al. (2010) ^42^ | Five times/week for 12 weeks; home-based | Three sets of 10 repetitions at 10 RM | hip abduction and adduction in a side-lying and standing position (with ankle cuff weights or elastic bands) |
| Bennell et al. (2014) ^43^ | Twelve times/12 weeks; supervised,  4 times/week; home-based | E1: NEXA  Three sets of 10 repetitions with 5 of 10 on a modified Borg CR-10 scale | 1. Forward and backward sliding or stepping  2. Sideways exercises  3. Functional hip muscle strengthening  4. Functional knee muscle strengthening  5. Step-up and down  6. Balance Quadriceps |
|  |  | E2: QS  Two or three sets of 10 repetitions at 10 RM | 1. Quads over a roll (inner range knee extension)  2. Knee extension in sitting  3. Knee extension with hold at 30° knee flexion  4. Straight leg raise  5. Outer range knee extension |
| Brenneman et al. (2018) ^44^ | Three or four times/week for 12 weeks; supervised | Seven on the Borg Perceived Exertion Scale | 1. Squats (different feet positions); lunges (different feet and arm positions); supported lunges; and transitions from standing to and from standing, sitting, and lying  2. Supine bridges and heel raises  3. Static stretching of major muscle groups of the lower limb. |
| Chang et al. (2016) ^45^ | Six to twelve sessions in 6 weeks; supervised  Five times/week for 6 weeks; home-based | Three sets with 10 repetitions (hold 10 sec)  Three repetitions (hold 90 sec) | 1. strengthening exercise for quadriceps, gluteus maximum, gluteus medius, and adductor magnus  2. muscle flexibility exercise for calf, hamstring, quadriceps femoris muscles  3. massage for tensor fasciae latae |

RM: repetition maximum; CR: category ratio; NEXA: Neuromuscular exercise; QS: quadriceps strengthening

(Continued)

| **Author (Year)** | **Frequency and Duration** | **Intensity** | **Contents of therapeutic exercise** |
| --- | --- | --- | --- |
| DeVita et al. (2018)^47^ | Three times/week for 3 months | Three sets of 10 repetitions with 60% of 3RM for first 2 weeks, 70% of 3RM for following 2 weeks and 85% of 3RM for the remaining 8 weeks | 1. warming up on a treadmill or stationary bicycle  2. leg extension, leg press and forward lunge exercise |
| Davis et al. (2019) ^46^ | Ten times in 28 days | daily adjustable progressive resistance exercise | 1. cycle ergometer and stretching (15 min)  2. progressive resistance exercise for hip abduction and knee extension and flexion (20 min)  3. balance progression (10 min)  Additional  ・exercise (described above) with TENS  ・exercise (described above) with placebo TENS  ・only exercise (described above) |
| Foroughi et al. (2011a) ^48^ | Three times/week for 6 months; supervised | E1: LI  Two sets of 8 repetitions at minimal resistance | 1. bilateral knee flexion, knee extension, leg press, and plantar-flexion |
|  |  | E2: PRT  Three of eight repetitions of 80% of 1RM (3% increments in resistance per session) | 1. unilateral knee extension  2. standing hip abduction and adduction  3. bilateral knee flexion, leg press, and plantar-flexion |
| Gaudreault et al. (2011) ^49^ | Twice/week for 12 weeks; supervised | Not detailed description | 1. isometric quadriceps contraction and half squat with a ball against a wall  2 .knee massage, patella mobilization, and passive stretching targeting the iliotibial band and the posterior muscle chain of the leg  3. stabilization on a ball, balance with one feet in front of the other, balance on a proprioceptive board with 3 points of support on the ground, balance on a proprioceptive board with only 1 point of support on the ground, and climbing and walking down 1 step as proprioceptive and balance exercise |

RM: repetition maximum; LI: low intensity; PRT: progressive resistance training; TENS: transcutaneous electrical nerve stimulation

(Continued)

| Author (Year) | Frequency and Duration | Intensity | Contents of therapeutic exercise |
| --- | --- | --- | --- |
| Henriksen et al. (2016)^50^ | Three times/week for 12 weeks; supervised | Not written | 1. warm-up phase (bicycle ergometer at moderate intensity)  2. circuit training program focusing on strength and coordination exercises of the trunk, hips and knees (with free weights, elastic rubber bands or body weight as resistance). |
| Holsgaard-Larsen et al. (2016)^51^ | Twice/ week for 8 weeks; supervised | 1. rather strenuous level  2-4. two sets of 12 repetitions | 1. warming up (ergometer cycling, treadmill, or stepper)  2. functional exercise consists of core stability/postural function, postural orientation, and lower-extremity muscle strength  3. proprioceptive part comprises three exercises, with the key elements being balance and functional stability  4. endurance strengthening part comprises three exercise circuits, with the key elements of postural and functional stability of the trunk and knee  5. cooling down |
| Hunt et al. (2013)^52^ | Five times a week 1,2,3,5 and 8; supervised  Four times/week for 10 weeks; home-based | Three sets of 10 repetitions | 1. standing and side lying hip abduction targeting hip abductors  2. standing lunges, mini-squats, and seated knee extension targeting quadriceps  3. knee flexion to 90° while maintaining standing balance on the contralateral limb targeting hamstrings |
| King et al. (2008)  ^53^ | Three times/week for 12 weeks | Three sets of 10 repetitions of 1RM for concentric exercise, three sets of fifteen repetitions at 60% of 1RM for reciprocal concentric isokinetic exercise | 1. concentric knee extension and flexion exercise at 60, 90, and 120 deg/s and reciprocal concentric isokinetic knee extension and flexion at 180 deg/s (with Biodex Multi-Joint System 3 dynamometer)  2. knee extensor and flexor stretches  3. stationary cycle ergometer (5 min) |
| Lim et al. (2008) ^54^ | Five times/week for 12 weeks; supervised home-based | Comfortably performed | 1. concentric exercise with ankle weight as long arc knee extension in the sitting position, inner range knee extension in supine lying or long sitting position, and straight leg raise in supine lying or elbow-supported supine lying position  2. isometric exercise with ankle weight and TheraBand™️ |
| Messier et al. (2013) ^55^ | Three times/week for 18 months | One or two sets of 10–12 repetitions | 1. aerobic walking (15min + 15min)  2. strength training (20min)  leg extension, leg press, seated leg curl, seated calf raise, compound row, and vertical chest or incline press  3. cool-down (10min) |

RM: repetition maximum
(Continued)

| Author (Year) | Frequency and Duration | Intensity | Contents of therapeutic exercise |
| --- | --- | --- | --- |
| Messier et al. (2021) ^56^ | Three times/week for 18 months | E1: LI  Three set of 15 repetitions at 30-40% 1RM  E2: HI  Three set of 75% and 80% of 1RM with 8 repetitions for first 2 weeks and week 3-4, respectively; 85% of 1RM with 6 repetitions for week 5-6; 90% of 1RM with 4 repetitions for week 7-8; established new 1RM after week 8 | 1. warm-up (5min)  2. exercise (6 lower body exercises and 4 upper body and core exercise)  hip abduction and adduction; leg curl, leg extension, and leg press; seated calf; compound row, vertical chest, lower back, and abdomen (40min)  3. cool-down (15min) |
| Pietrosimone et al. (2011) ^60^ | Ten times in 28 days | Daily adjustable progressive resistance exercise | 1. cycle ergometer and stretching  2. progressive resistance exercise for hip abduction and knee extension and flexion  3. balance progression  Additional  TENS group: exercise (described above) with TENS  Placebo TENS group: exercise (described above) with placebo TENS  Control group: only exercise (described above) |
| Sled et al. (2010) ^57^ | Three or four times/week for 8 weeks; home-based | At a level that does not cause fatigue after 20 times repetitions | side-lying resistive exercises for the hip abductor muscles; standing single-leg stabilization exercises; and single-leg standing exercise off the side of a 10-cm step |
| Thorp et al. (2010) ^58^ | Three times for the first 2 weeks, once/week for the next 2 weeks; supervised  4 times/week for the first 2 weeks, 6 times/week for the next 2 weeks; home-based | Three repetitions, 15 seconds hold each repetition  Three sets, 10 repetitions each | 1. isometric exercise targeting on the hamstrings in the supine or long sitting position, quadriceps in the sitting position, and hip abductors in a side lying position  2. concentric exercise targeted at the hamstrings in both standing and prone positions, quadriceps in the standing position, and hip abductors in a side lying position, standing on one leg  3. stretching exercise targeted at hip abductors  Isometric exercise (home exercise) |
| Thorstensson et al. (2007) ^59^ | Twice/week for 8 weeks; supervised | Three set of 15 repetitions  Three set of 60 seconds for rebounder | 1. warming up (ergometer cycle or treadmill)  2. sit-ups, hip lift and lunging forward  3. proprioceptive task, knee bendings, and rebounder exercise  4. knee control on slippery surface, during step up and down, and during straddle-legged body weight transfer  5. pulley exercises in four directions: hip extension, abduction, flexion and adduction  6. stretching exercises for muscles triceps surae, quadriceps, hamstrings, and iliopsoas were performed. |

RM: repetition maximum; LI: low intensity; HI: high intensity; TENS: transcutaneous electrical nerve stimulation

**Supplementary Table 4.** List of variables for biomechanical loads and potential moderator and mediator candidates

| Author (Year) | Biomechanical loads | | | | Potential  moderators | | Potential mediators | | | | | | | | | |
| --- | --- | --- | --- | --- | --- | --- | --- | --- | --- | --- | --- | --- | --- | --- | --- | --- |
|  | First peak KAM | Peak KFM | Knee joint compression force | Co-contraction Index | OA severity | Joint deformity | Hip flexor muscle strength | Hip extensor muscle strength | Hip abductor muscle strength | Hip adductor muscle strength | Hip internal rotator muscle strength | Hip external rotator muscle strength | Knee extensor muscle strength | Knee flexor muscle strength | Pain | Walking speed |
| Al-Khlaifat (2016) ^41^ | **●** |  |  | **●** |  |  |  |  | **●**^¶^ |  |  |  | **●**^¶^ | **●**^¶^ | **●**^‡‡^ |  |
| Bennell (2010) ^42^ | **●**^*^ | **●**^*^ |  |  | **●** | **●**^*^ | **●** | **●** | **●** | **●** | **●** | **●** | **●** |  | **●** |  |
| Bennell (2014) ^43^ | **●**^*^ | **●**^*^ |  |  | **●** | **●**^*^ |  | **●** | **●** |  | **●** | **●** | **●** | **●** | **●** | **●** |
| Brenneman (2015) ^44^ | **●**^†^ |  |  |  |  |  |  |  |  |  |  |  | **●** | **●** | **●**^‡‡^ |  |
| Chang (2016) ^45^ | **●** | **●** |  |  |  |  |  |  |  |  |  |  |  |  |  | **●** |
| Davis (2019)^46^ |  | **●**^§^ |  |  | **●** |  |  |  |  |  |  |  | **●** |  | **●** | **●** |
| DeVita (2018)^47^ |  |  | **●** |  | **●** |  |  |  |  |  |  |  | **●** |  | **●** | **●** |
| Foroughi (2011a) ^48^ | **●** |  |  |  | **●**^\|\|^ |  |  |  |  |  |  |  | **●** |  | **●** | **●** |
| Gaudreault (2011) ^49^ | **●** | **●** |  |  | **●** |  |  |  |  |  |  |  | **●** | **●** | **●** |  |
| Henriksen (2016) ^50^ | **●**^*^ | **●**^*^ |  |  | **●**^*^ | **●**^*^ |  |  |  |  |  |  |  |  | **●**^*,‡‡^ | **●**^*^ |
| Holsgaard-Larsen (2016) ^51^ | **●**^‡^ |  |  |  | **●** |  |  |  |  |  |  |  |  |  | **●**^‡‡^ |  |
| Hunt (2013)^52^ | **●**^*^ |  |  |  | **●**^*^ | **●**^*^ |  |  | **●** |  |  |  | **●** | **●** |  | **●** |
| King (2008) ^53^ | **●**^†^ |  |  |  | **●** | **●** |  |  |  |  |  |  | **●**^††^ | **●**^††^ | **●**^‡‡^ | **●** |
| Lim (2008) ^54^ | **●**^*^ | **●**^*^ |  |  | **●** | **●** |  |  |  |  |  |  | **●** |  | **●** |  |
| Messier (2013) ^55^ |  |  | **●** |  | **●** |  |  |  |  |  |  |  |  |  | **●** | **●** |
| Messier (2021) ^56^ |  |  | **●** |  | **●** |  |  |  | **●**^**^ |  |  |  | **●**^**^ |  | **●**^§§^ |  |
| Pietrosimone (2010) ^60^ |  | **●** |  |  | **●** |  |  |  |  |  |  |  |  |  |  | **●** |
| Sled (2010) ^57^ | **●** |  |  |  | **●** |  |  |  | **●** |  |  |  |  |  | **●** |  |
| Thorp (2010) ^58^ | **●**^†^ |  |  |  |  |  |  |  | **●**^§^ |  |  |  | **●** | **●** | **●**^§§^ |  |
| Thorstensson (2007) ^59^ | **●**^§^ |  |  |  | **●** | **●** |  |  |  |  |  |  |  |  |  |  |

KAM: knee adduction moment; KFM: knee flexion moment; OA: osteoarthritis

†* Unpublished data; authors provided variables as a favor following our inquiry.

*† Peak KAM value was included in this systematic review as the first peak KAM.

§‡ Postexercise value was calculated from the mean difference and baseline value.

‡§ Variable was extracted a figure.

|| Not included in the regression analysis because the modified Outerbridge classification system with magnetic resonance imaging grading was used to evaluate OA severity (most studies used the Kellgren-Lawrence grade).

¶ Not included in this systematic review due to the unreliability of the results.

** Not included for regression-analysis because variables were adjusted or sex, baseline BMI, and baseline outcome values

†† Not included in the regression analysis because isokinetic muscle strength was measured (most studies measured isometric muscle strength)

‡‡ Not included in the regression analysis because the Knee injury and Osteoarthritis Outcome Score was used (Most studies used the Western Ontario and McMaster Universities Osteoarthritis Index pain subscale).

**Supplementary Table 5.** Risk of bias in randomized controlled trials

| Author (year) | Eligibility criteria | Random allocation | Concealed allocation | Baseline comparability | Blind subjects | Blind therapists | Blind assessors | Adequate follow-up | Intention-to-treatment analysis | Between-group comparisons | Point estimates and variability | Score |
| --- | --- | --- | --- | --- | --- | --- | --- | --- | --- | --- | --- | --- |
| Bennell et al. (2010) ^42^ | Y | Y | Y | Y | N | N | Y | Y | Y | Y | Y | 8 |
| Bennell et al. (2014) ^43^ | Y | Y | Y | Y | N | N | N | Y | Y | Y | Y | 7 |
| DeVita et al. (2018)^47^ | N | Y | N | Y | N | N | N | Y | Y | Y | Y | 6 |
| Foroughi et al. (2011a) ^48^ | N | Y | Y | N | N | N | N | N | Y | Y | Y | 5 |
| Henriksen et al. (2016) ^50^ | Y | Y | Y | Y | N | N | Y | N | N | Y | Y | 6 |
| Holsgaard-Larsen et al. (2016) ^51^ | Y | Y | Y | Y | Y | N | N | Y | Y | Y | Y | 8 |
| Hunt et al. (2013)^52^ | Y | Y | N | Y | N | N | Y | Y | Y | Y | Y | 7 |
| Lim et al. (2008) ^54^ | Y | Y | Y | Y | N | N | Y | Y | Y | Y | Y | 8 |
| Messier et al. (2013) ^55^ | Y | Y | N | Y | Y | N | N | Y | Y | Y | Y | 7 |
| Messier et al. (2021) ^56^ | Y | Y | N | Y | N | N | Y | N | Y | Y | Y | 6 |
| Pietrosimone et al. (2010) ^60^ | Y | Y | N | Y | Y | N | Y | Y | N | Y | Y | 7 |

**Supplementary Table 6.** Risk of bias in non-randomized controlled trials

|  | #1 | Was the study question or objective clearly stated? | | | | | | | | | | | | | |
| --- | --- | --- | --- | --- | --- | --- | --- | --- | --- | --- | --- | --- | --- | --- | --- |
|  | #2 | Were eligibility/selection criteria for the study population prespecified and clearly described? | | | | | | | | | | | | | |
|  | #3 | Were the participants in the study representative of those who would be eligible for the test/service/intervention in the general or clinical population of interest? | | | | | | | | | | | | | |
|  | #4 | Were all eligible participants that met the prespecified entry criteria enrolled? | | | | | | | | | | | | | |
|  | #5 | Was the sample size sufficiently large to provide confidence in the findings? | | | | | | | | | | | | | |
|  | #6 | Was the test/service/intervention clearly described and delivered consistently across the study population? | | | | | | | | | | | | | |
|  | #7 | Were the outcome measures prespecified, clearly defined, valid, reliable, and assessed consistently across all study participants? | | | | | | | | | | | | | |
|  | #8 | Were the people assessing the outcomes blinded to the participants' exposures/interventions? | | | | | | | | | | | | | |
|  | #9 | Was the loss to follow-up after baseline 20% or less? Were those lost to follow-up accounted for in the analysis? | | | | | | | | | | | | | |
|  | #10 | Did the statistical methods examine changes in outcome measures from before to after the intervention? Were statistical tests done that provided p values for the pre-to-post changes? | | | | | | | | | | | | | |
|  | #11 | Were outcome measures of interest taken multiple times before the intervention and multiple times after the intervention (i.e., did they use an interrupted time-series design)? | | | | | | | | | | | | | |
|  | #12 | If the intervention was conducted at a group level (e.g., a whole hospital, a community, etc.) did the statistical analysis take into account the use of individual-level data to determine effects at the group level? | | | | | | | | | | | | | |
| Author (year) | | | 1 | 2 | 3 | 4 | 5 | 6 | 7 | 8 | 8 | 10 | 11 | 12 | Score |
| Al-Khlaifat et al. (2016) ^41^ | | | Y | Y | Y | CD | CD | Y | Y | CD | N | Y | CD | N | 6 |
| Brenneman et al. (2015) ^44^ | | | Y | Y | Y | CD | CD | Y | Y | N | Y | Y | CD | CD | 7 |
| Chang et al. (2016) ^45^ | | | Y | Y | Y | CD | CD | Y | Y | CD | Y | Y | CD | CD | 7 |
| Davis et al. (2019)^46^ | | | Y | Y | Y | CD | CD | Y | Y | CD | CD | Y | CD | CD | 6 |
| Gaudreault et al. (2011) ^49^ | | | Y | Y | Y | CD | CD | CD | Y | CD | CD | Y | CD | CD | 5 |
| King et al. (2008) ^53^ | | | Y | Y | N | CD | CD | Y | Y | CD | CD | Y | CD | CD | 5 |
| Sled et al. (2010) ^57^ | | | Y | Y | Y | CD | CD | Y | Y | CD | Y | Y | CD | CD | 7 |
| Thorp et al. (2010) ^58^ | | | Y | Y | CD | CD | CD | CD | Y | CD | Y | Y | CD | CD | 5 |
| Thorstensson et al. (2007) ^59^ | | | Y | Y | Y | CD | CD | Y | Y | CD | Y | Y | CD | CD | 7 |

**Supplementary Figure 1.** Standardized mean difference (SMD) and 95% confidence interval (CI) for muscle strength of knee extensor (A) and knee flexor (B) in each group.

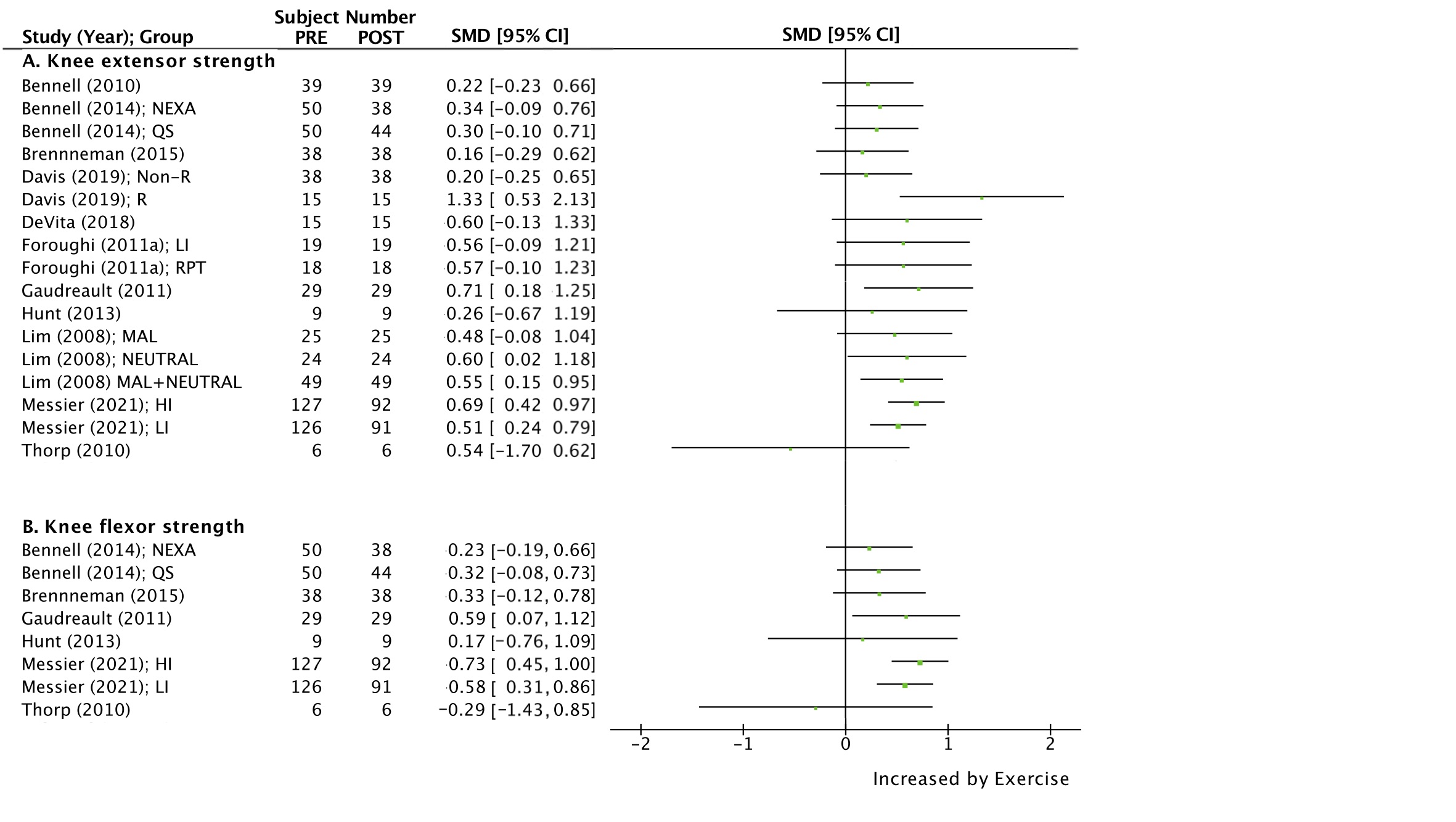

**Supplementary Figure 2.** Standardized mean difference (SMD) and 95% confidence interval (CI) for muscle strength of hip extensor (A), hip flexor (B), hip abductor (C), hip adductor (D), hip external rotator (E), and hip internal rotator (F) in each group.

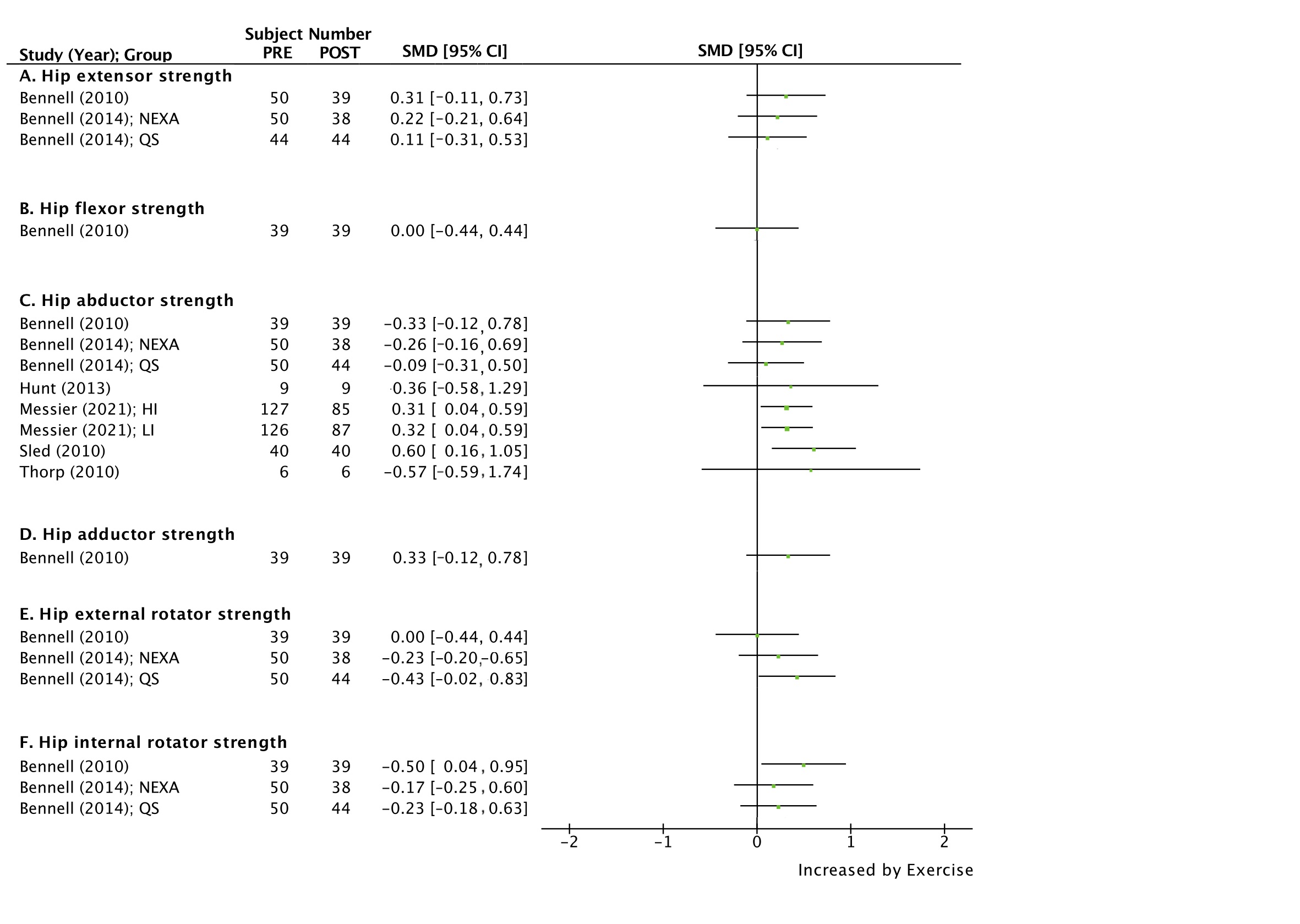

**Supplementary Figure 3.** Standardized mean difference (SMD) and 95% confidence interval (CI) for WOMAC pain (A) and walking speed (B) in each group.

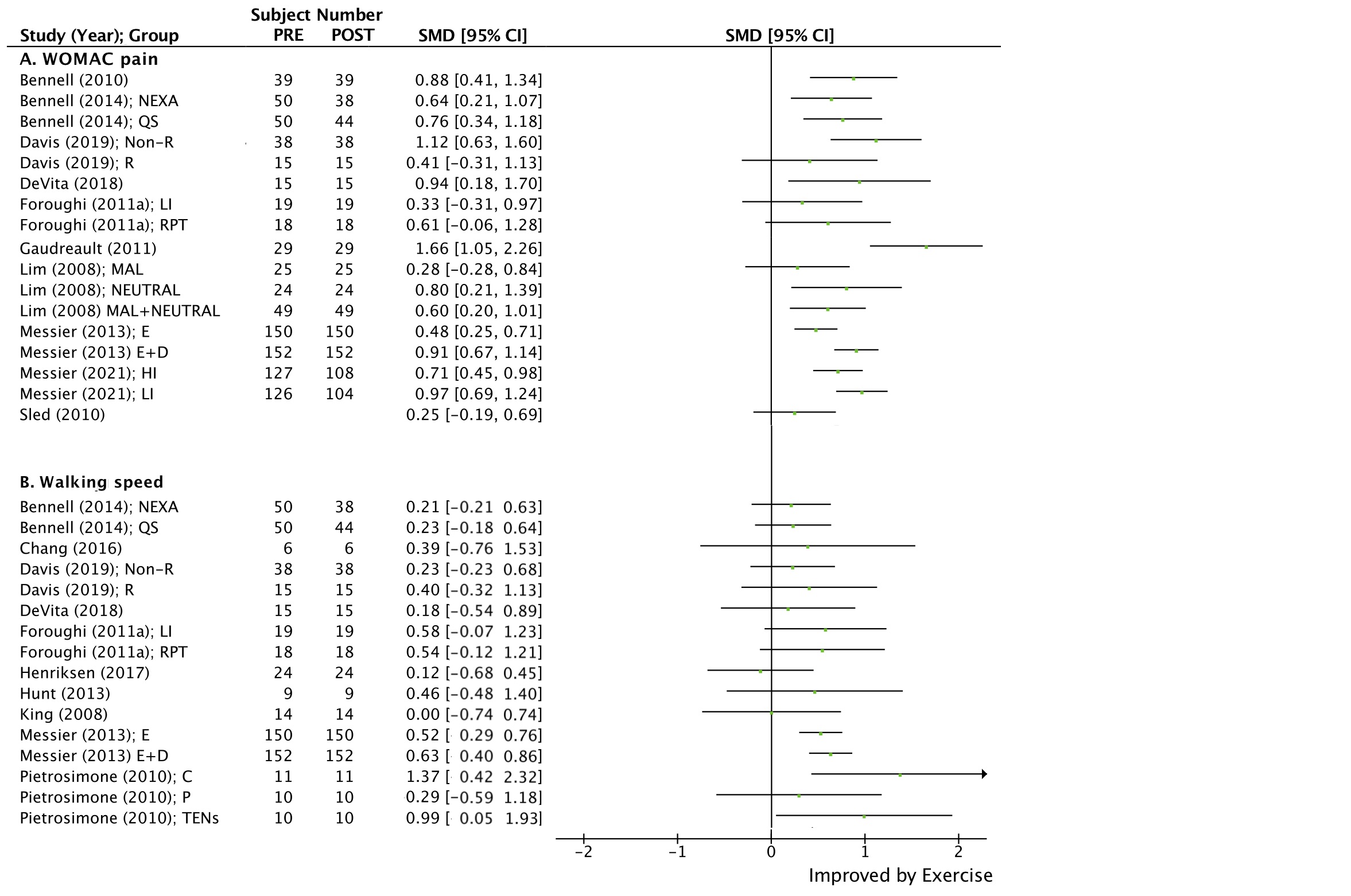

**Supplementary Figure 4.** Relationship between standardized mean difference (SMD) for first peak knee adduction moment (KAM) or first peak knee flexion moment (KFM) and potential moderator candidates.

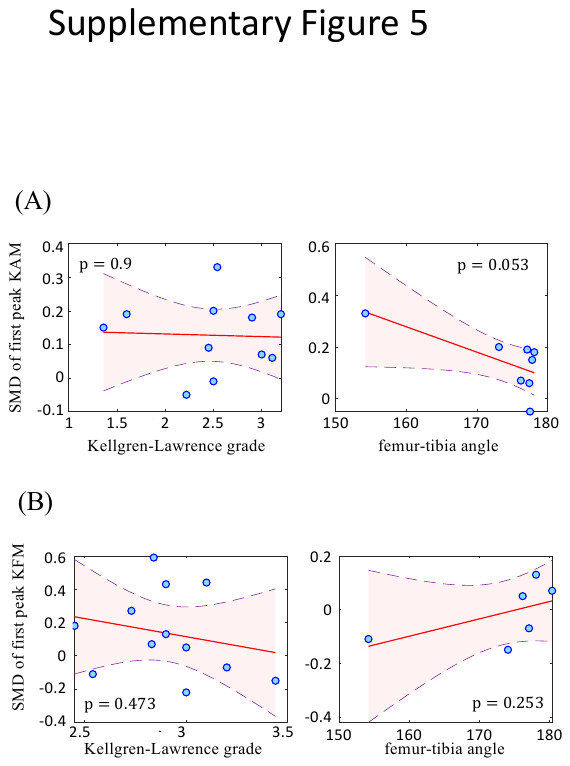

**Supplementary Figure 5.** Relationship between standardized mean difference (SMD) for first peak knee adduction moment (KAM), first peak knee flexion moment (KFM), or maximal knee joint compression force (KCF) and potential mediator candidates.

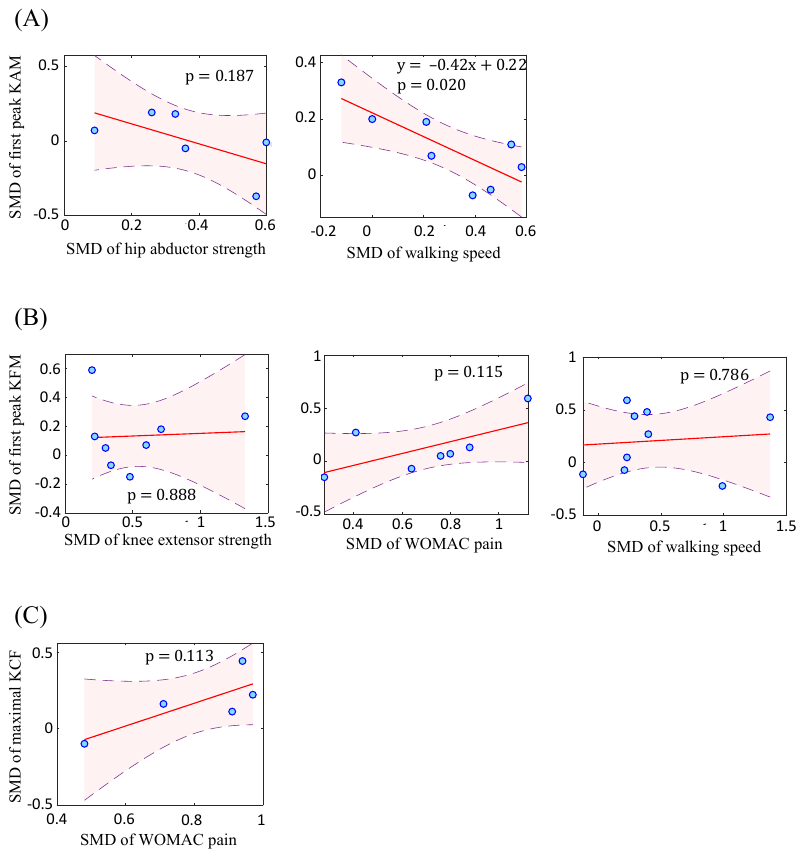

**Supplementary Figure 6.** Funnel plot representing publication bias for the effect of exercise intervention on peak knee adduction moment (A), knee adduction moment first peak (B), and knee flexion moment (C).

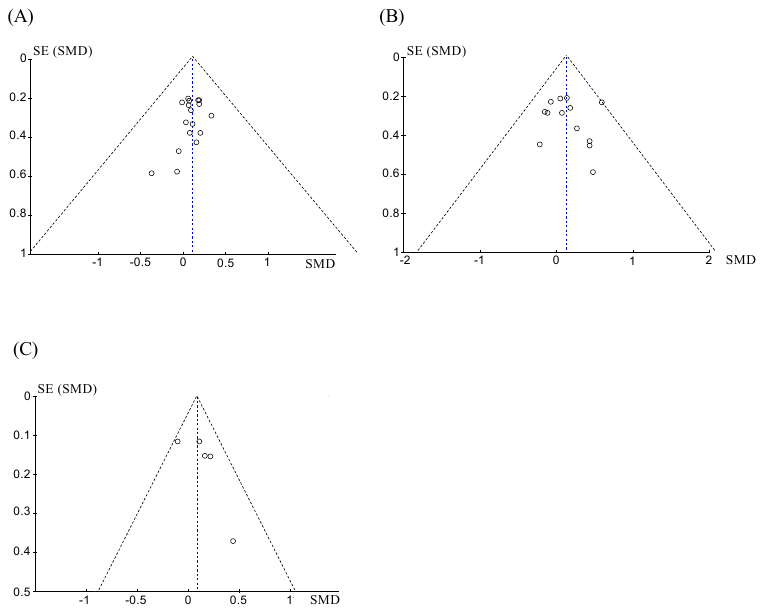
